# Mapping tumor heterogeneity: ex-vivo drug responses in matched primary and metastatic tumors using a 3D functional precision platform

**DOI:** 10.64898/2026.09.22.26357781

**Authors:** Rajeshwar Nitiyanandan, Ivan Trus, Chiara Maestri, Ricardo J. Parker, William Cance, Brigitte Apfel, Christian Apfel

**Affiliations:** SageMedic Corp, Redwood City, CA, USA; International Institute of Molecular and Cell Biology, Warsaw, Poland; National University, San Diego, CA, USA; University of Arizona, Tucson, AZ, USA; University of California San Francisco, San Francisco, CA, USA

## Abstract

**Introduction:** Tumors are known to develop genomic heterogeneity over time, particularly in metastatic disease, raising concerns that a single-site biopsy may be insufficient to accurately predict response to medications.

**Methods:** We used our 3D microtumor functional profiling platform that preserves key features of the native tumor microenvironment. We tested (i) six split samples from primary tumors and (ii) seven samples from primary tumors (ovarian, colon, and breast cancer) matched with their locoregional or distant metastases. Fresh tissue specimens were processed within 24 hours after surgery and treated with NCCN guideline-recommended chemo or targeted therapies at concentrations over four orders of magnitude. Cytotoxicity was evaluated after three days to generate dose–response curves. The response correlations between the split sample or between the two matched sites were quantified using the concordance correlation coefficient (ρc) for the drug efficacy metrics.

**Results:** We compared a total of 275 drug responses. The six split samples resulted in a strong correlation between the two tumor areas (ρc=0.95, p<0.001), supporting high assay reproducibility across distinct regions of the same tumor. Locoregional metastases were highly correlated with their primaries (ovarian–omentum ρc=0.97, p<0.001; ovarian–liver capsule ρc=0.93, p<0.001; colon–peritoneum ρc=0.91, p<0.001). A lesser correlation was observed with lymphogenic metastases (breast–axilla ρc=0.87, p<0.001), and the correlations of hematogenic metastases were even lower (colon–lung ρc=0.69 p<0.001; ovarian–soft tissue/colon ρc=0.58, p<0.001). No correlation was observed among specimens that were resistant to all tested medications.

**Conclusion:** Tumor drug responses were highly preserved in locoregional metastases, whereas somewhat greater divergence was observed in lymphogenic or hematogenic metastases. Single-site functional profiling may remain informative for identifying treatment resistance and might reduce the risk of ineffective therapy selection in metastatic disease.

---

Intra-tumoral heterogeneity is a major biological driver of therapeutic resistance because clinically relevant sensitivities and resistant subclones can coexist within the same patient. This tumor heterogeneity can undermine treatment selection when decisions are based on a limited tissue sample, particularly when a single biopsy is assumed to represent the broader disease^1–4^. This problem is fundamentally spatial: different regions within a primary tumor can harbor distinct clonal architectures and microenvironmental states, and these differences can translate into variable drug vulnerabilities^3,5^. Classic multiregional sampling studies demonstrate that single-site biopsies may under-represent the diversity present within a tumor mass^1,6^.

A parallel and clinically urgent sampling question arises in metastatic disease: whether the most informative specimen for guiding therapy is the primary tumor, a local metastatic deposit, a lymph node metastasis, or a distant hematogenous metastasis^7,8^. These routes of dissemination are biologically distinct and expose tumor cells to different selective pressures and stromal/immune niches, plausibly shaping functional drug response. Lymphatic spread is often an early conduit for dissemination in carcinomas, whereas hematogenous spread is more commonly associated with distant organ metastasis; both have been linked to differences in metastatic progression and tumor–microenvironment interactions^8^. In addition, several tumor types (including ovarian and gastrointestinal malignancies) can exhibit local or cavity-associated spread (e.g., peritoneal/omental involvement), where proximity and shared environments may preserve aspects of primary-tumor biology, yet this should not be assumed without empirical testing.

Functional precision oncology approaches aim to complement molecular profiling by directly measuring drug response in living patient-derived material^9–12^. Ex vivo drug testing platforms including 3D microtumor and organotypic models seek to retain elements of native architecture, cell–cell interactions, and microenvironmental context that can influence treatment sensitivity^9,13^. However, functional assays are also susceptible to sampling bias: the measured response can depend on where the tissue was obtained, both across lesions (primary vs metastasis) and within a single primary tumor (different regions of the same mass). As functional profiling moves toward clinical implementation, defining how biopsy location and dissemination route affect observed drug response becomes essential for interpreting results and for designing practical sampling strategies^8^,^14–16^.

In this study, we use a CLIA-validated 3D microtumor functional profiling assay to evaluate how drug-response profiles vary (i) among biopsies from adjacent regions of a primary tumor and (ii) between primary tumors and their matched metastases arising via local, lymphogenic, or hematogenic dissemination^17–20^.

## Results

### Intratumoral spatial heterogeneity

To determine whether sampling from different regions within the same tumor influences functional profiling results, we evaluated multiple biopsies obtained from distinct intratumoral locations (Fig. 1). Correlation between spatially separated regions varied by tumor type. Strong agreement (ρc=0.95, p<0.001) was observed in 6 samples with 110 pairwise drug comparisons (Fig. 2), including breast cancer (ρc=0.89, p<0.001) and colon tumors (ρc=0.95, p<0.001; ρc=0.92, p<0.001; ρc=0.89, p<0.001). No correlation was determined when both regions were broadly resistant across the drug panel.

**Fig. 1.**
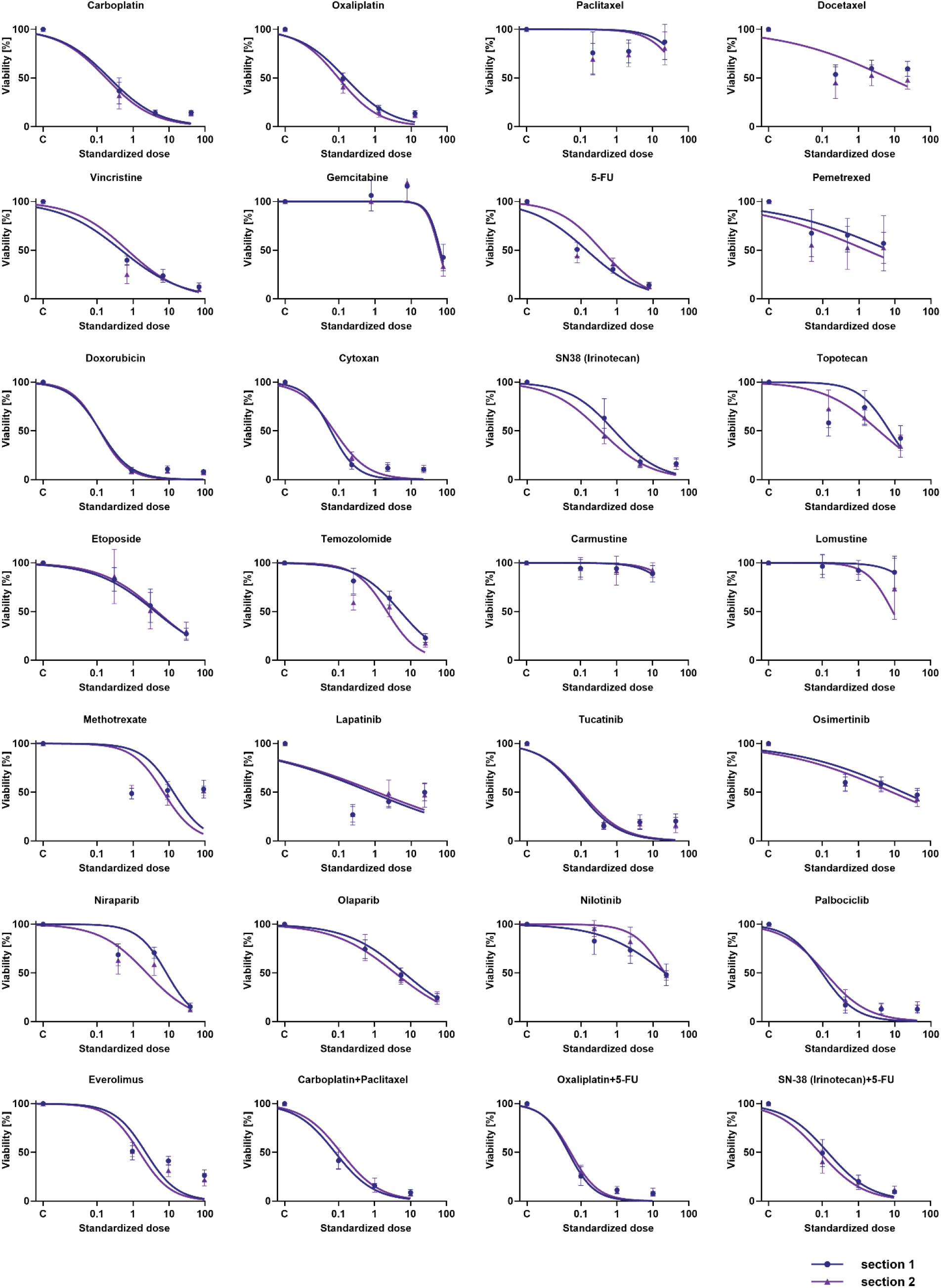
Comparison of dose–response curves from several chemo or targeted therapies of two biopsies obtained at different locations within the same primary breast tumor. Whiskers represent 95% confidence intervals.

**Fig. 2.**
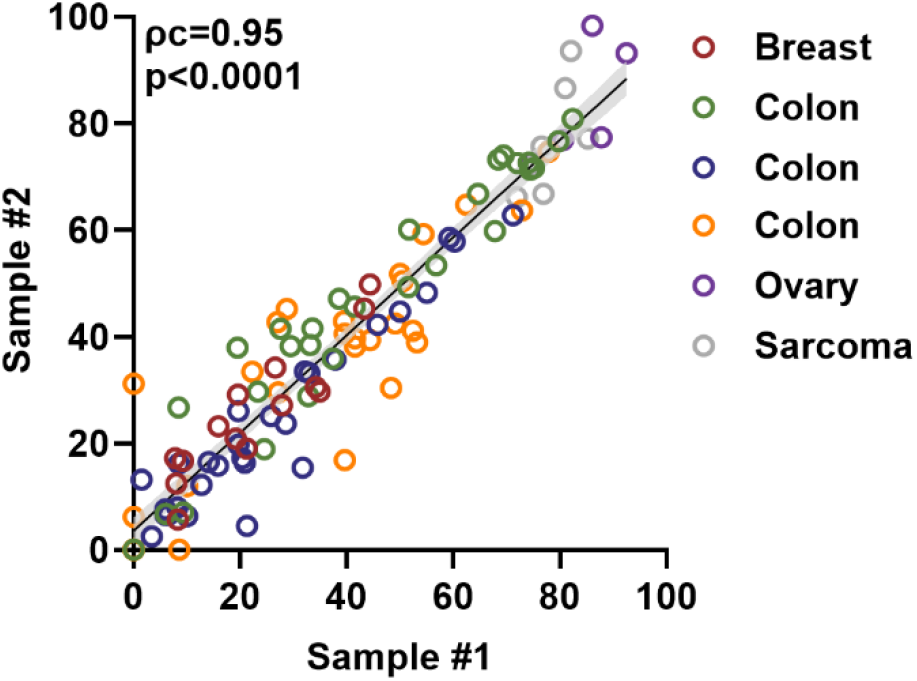
Scatter plot showing correlation of standardized effective dose between two spatially distinct tumor samples (Sample #1 vs #2). Each circle represents a chemo or targeted therapy tested in a single patient; colors denote individual patients (n = 6). Individual correlation plots are shown in Supplementary Fig. S1.

### Overview of matched primary–metastatic drug response profiling

Following validation of assay reproducibility using spatially separated primary samples, matched primary–metastatic comparisons were analyzed (Figs. 3 and 4). Pairwise drug comparisons (n = 165) were generated from seven patients with synchronous matched primary and metastatic tumors across Local (locoregional) metastases generally exhibited the highest correlation with their corresponding primary tumors (Fig. 5, Supplementary Fig. S2). An ovarian cancer primary–omentum metastasis pair demonstrated near-identical drug-response profiles (ρc=0.97, p<0.001). Similarly, ovarian cancer metastasis to the liver capsule showed strong correlation with the primary tumor (ρc=0.93, p<0.001). A colon cancer primary–peritoneal metastasis pair also exhibited high correlation (ρc=0.91, ovarian, colon, and breast cancers (Fig. 5). Drug responses were quantified using DSS (Drug Sensitivity Score) metrics at standardized drug concentrations. In general, across all matched comparisons, a strong correlation was observed between primary and local/lymphogenic metastases (ρc=0.93, p<0.001) though there was a significantly lower correlation with distant metastases (ρc=0.67, p<0.001). A second ovarian cancer primary–omentum metastasis pair showed similarly flat dose–response curves across the drug panel, consistent with broad resistance; however, correlation metrics were low, likely reflecting the compressed response dynamic range associated with near-uniform non-response. Therefore, here ρc values should be interpreted alongside the underlying dose–response shapes and response dynamic range.

**Fig. 3.**
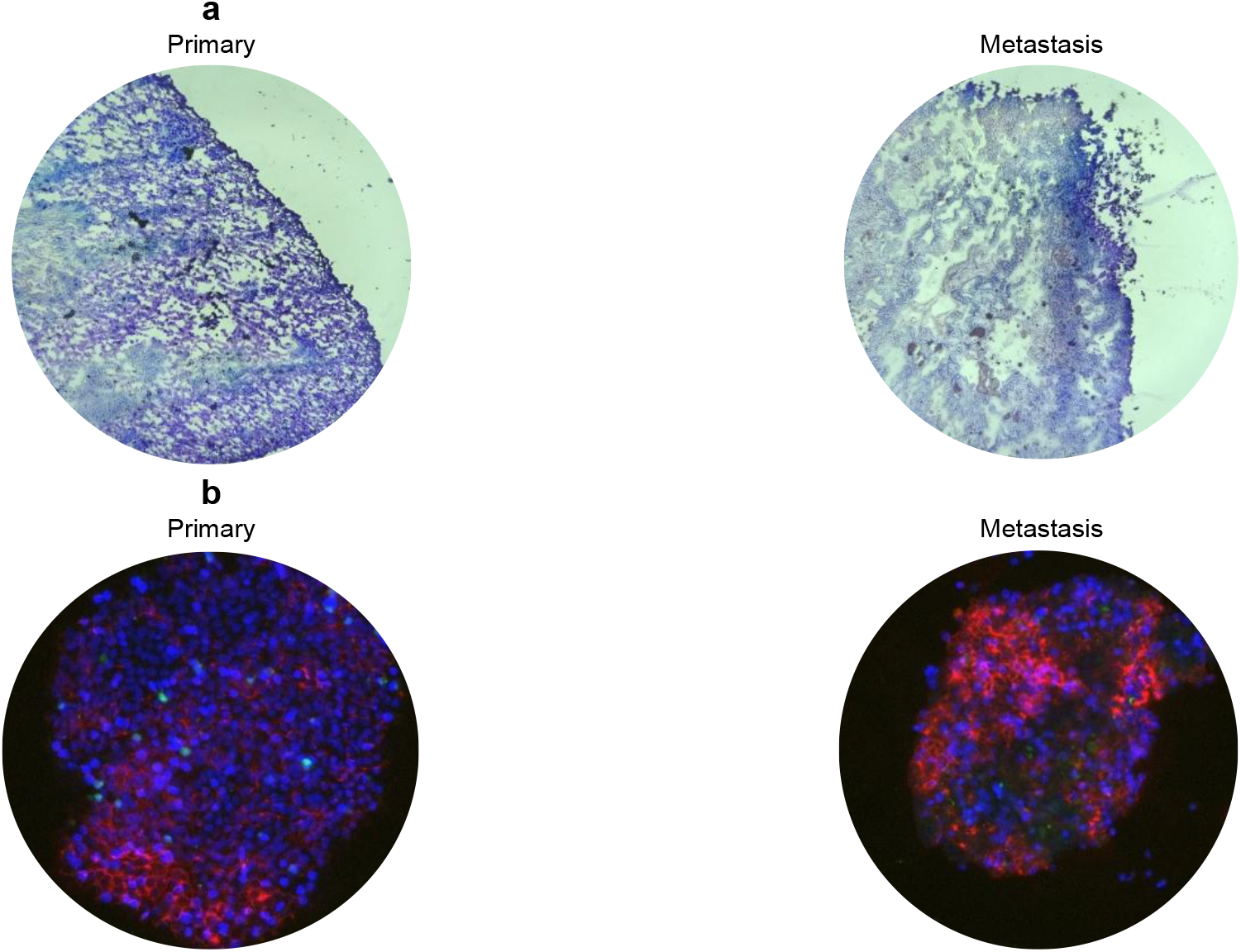
Example of a colon primary tumor and its metastasis: (**a**) H&E staining and (**b**) immunofluorescent staining with the tumor marker EpCAM (red), the leukocyte marker CD45 (green), and the nuclear stain Hoechst 33342 (blue).

**Fig. 4.**
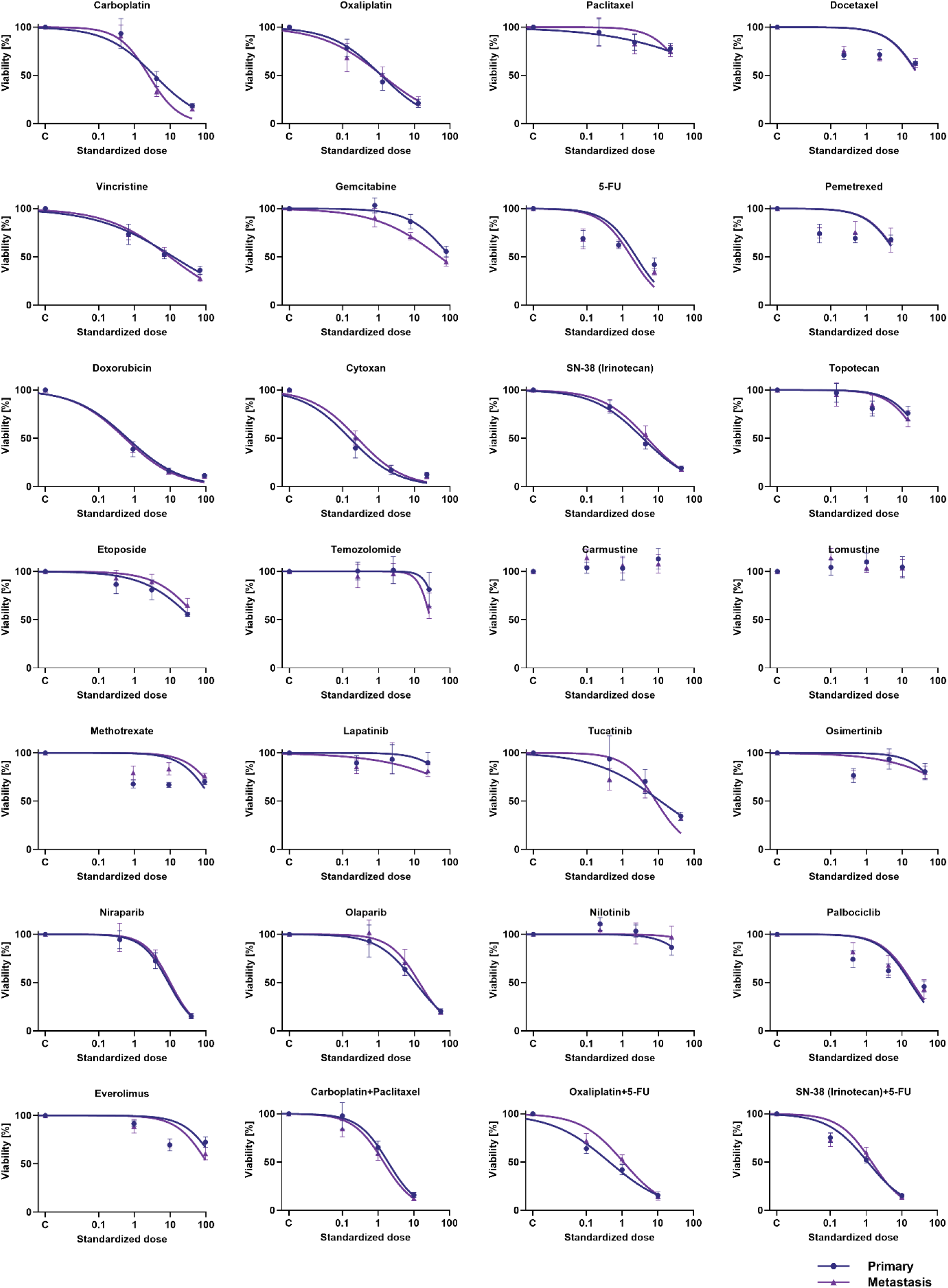
Comparison of dose–response curves of an ovarian primary tumor with its regional metastasis (omental). Whiskers represent 95% confidence intervals.

**Fig. 5.**
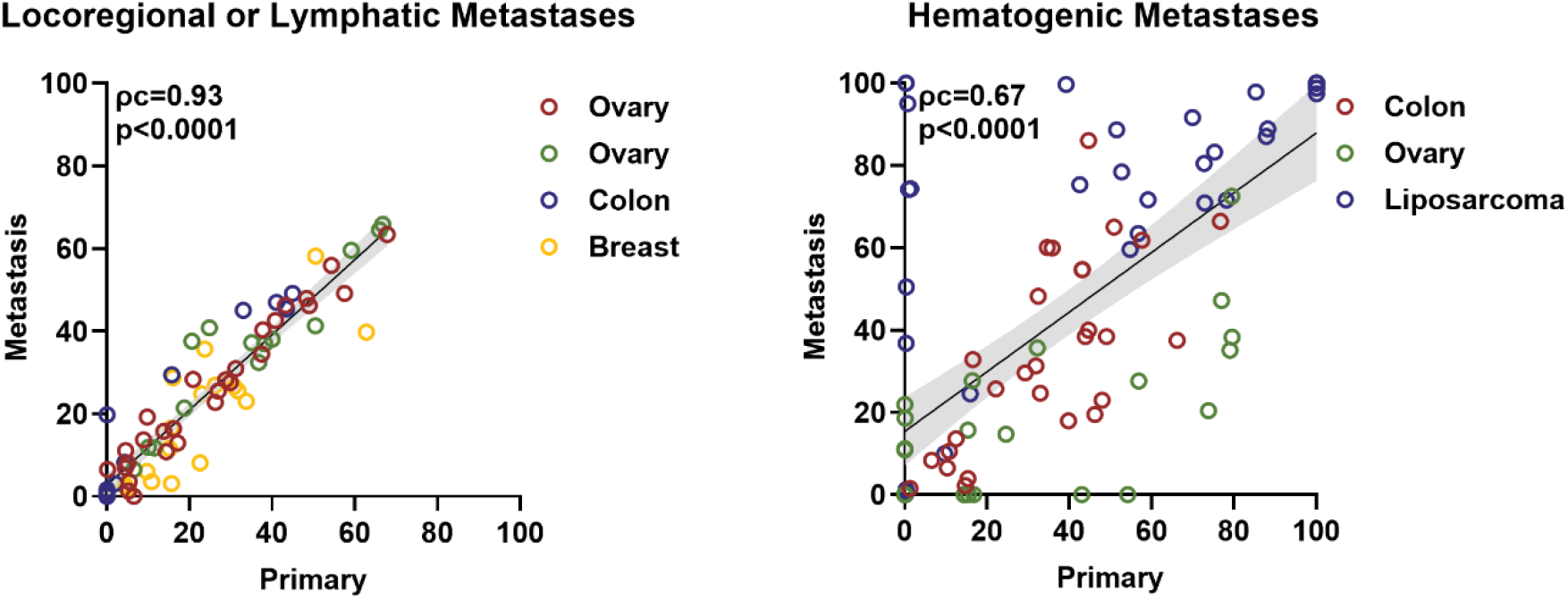
Correlation charts of the effective standardized dose in seven pairs of a primary tumor and their metastasis. A stronger correlation (ρc = 0.93) was observed for locoregional and lymphatic metastases; a weaker correlation was found for hematogenic metastases (ρc = 0.67). Individual correlation plots are shown in Supplementary Fig. S2.

Lymphogenic metastases displayed intermediate and variable correlation with primary tumors (Supplementary Fig. S2). A breast cancer primary–axillary lymph node metastasis pair demonstrated moderate agreement in DSS (ρc=0.89, p<0.001).

Hematogenic metastases consistently showed lower correlation with primary tumors compared with locoregional disease (Fig. 5, Supplementary Fig. S2). A colon cancer primary–lung metastasis pair demonstrated modest correlation (ρc=0.69, p<0.001). An ovarian cancer case with hematogenic spread to soft tissue and colon also showed reduced correlation (ρc=0.58, p<0.001), and a liposarcoma case with hematogenic spread to two different locations in the lungs showed reduced correlation (ρc=0.54, p<0.001).

Consistent with the correlation analysis, Bland–Altman plots showed close agreement between primary tumor samples, and locoregional and lymphatic metastases, whereas hematogenic metastases showed broader limits of agreement (Supplementary Fig. S3).

## Discussion

Tumor heterogeneity complicates therapeutic decision-making by introducing uncertainty about whether a single biopsy accurately reflects clinically relevant drug sensitivities^1–5^. In this study, spatially separated biopsies from primary tumors were first evaluated to establish the robustness and reproducibility of a rapid 3D microtumor functional profiling assay. Across multiple tumor types, highly reproducible drug-response profiles were observed, providing confidence that the platform can reliably detect functional similarities and differences across independently processed samples^9,21^. This technical validation is critical for interpreting subsequent comparisons between primary tumors and metastatic lesions.

When applied to matched primary and metastatic specimens, functional drug-response correlation was dependent on the anatomical site and route of metastatic dissemination. Locoregional metastases, including omental, peritoneal, and liver capsule involvement, most frequently exhibited high correlation with their corresponding primary tumor DSS metrics. These findings indicate that functional drug sensitivity and resistance spectra of local metastases of primary tumors are preserved in most cases. From a clinical perspective, this suggests that functional profiling of either the primary tumor or a locoregional metastasis may be sufficient to generate representative drug-response profiling, potentially reducing the need for multiple invasive biopsies in these settings.

In contrast, lymphogenic and hematogenic metastases showed greater variability and overall lower correlation with primary tumors. While some lymph node metastases retained moderate similarity to the primary tumor, others diverged markedly, indicating that lymphatic dissemination does not consistently preserve primary-tumor drug-response behavior. Hematogenic metastases, particularly those involving distant organs such as lungs or soft tissue, demonstrated the weakest correlation overall. These observations suggest that reliance on primary-tumor profiling alone may be insufficient in certain metastatic contexts and that functional profiling of metastatic lesions may provide additional, clinically relevant information.

## Conclusion

Functional drug responses were largely conserved across regions of the same tumor and between primary tumors and locoregional metastases, whereas lymphatic and hematogenous metastases showed greater divergence. These findings suggest that profiling a single accessible lesion may often provide clinically useful information, particularly for identifying likely resistance, while testing a metastatic site may add value when distant disease is present. Thus, rapid 3D microtumor profiling could support treatment prioritization while reducing the risk of selecting ineffective therapies.

## Methods

### Sample collection

Biopsies obtained from adjacent regions of the same tumor, as well as matched primary and metastatic tumor specimens (ovarian, colon, and breast cancers; metastases including omentum, liver capsule, peritoneum, lung, axilla, soft tissue, and ascites), were collected, cooled, and shipped overnight as independent specimens to SageMedic Corp. (Redwood City, California, USA) using a proprietary transport medium to preserve sample integrity during transit, and were processed immediately upon receipt (within 24 hours of collection) to generate patient-derived microtumors that preserve native architecture and key features of the patient’s tumor microenvironment, as previously described^22^.This study analyzed clinical specimens and associated data obtained through SAGE Oncotest testing and/or approved secondary research use. The secondary-use research was conducted under the program titled “Secondary Research Analyses of SAGE Oncotest Results and Associated Clinical Data,” which Compassionate Access IRB reviewed and classified as exempt pursuant to 45 CFR §46.104(d)(4)(iii) (IORG0012307; CAIRB-26-0901). Research activities were limited to evaluating existing assay results and associated clinical data. No patient interventions, treatment assignments, or additional patient procedures were undertaken specifically for the research.

### Sample plating

Tissue fragments from solid tissues and enriched malignant cell aggregates from ascites were plated in a 3D format on 384-well plates to generate patient-derived microtumors and incubated overnight at 37°C in a humidified incubator with 5% CO_2_ to allow recovery and stabilization prior to drug exposure.

### Drug treatment

The next day (day 1), microtumors were treated with a patient-specific panel of NCCN-recommended and mechanistically relevant therapies, with 6–24 drugs (single agents and/or combinations) tested per patient. Representative agents among others included platinum-based compounds, taxanes, 5-fluorouracil, irinotecan, gemcitabine, PARP inhibitors (olaparib, rucaparib) and cyclophosphamide. Drugs were tested across a wide concentration range that included physiologically relevant ranges.

### Drug cytotoxicity assay

On day 4, a final readout was performed to measure drug cytotoxic effects, as described elsewhere^23^. For structural immunohistochemical characterization and imaging, 3D microtumors were counterstained with Hoechst 33342 (Cat. #40044; Biotium, California, USA). Dose–response curves were generated.

### H&E staining

A portion of each tissue sample was frozen in optimal cutting temperature (OCT) compound (Cat. #4583; Sakura Finetek, California, USA), and 8 µm sections were generated using a cryotome (Shandon, Thermo Fisher Scientific, Massachusetts, USA). The cryosections obtained were mounted onto glass slides and dried at 37°C for 30 minutes before staining. After hydrating in distilled water for 2 minutes, cryosections were stained using the hematoxylin and eosin (H&E) protocol from Vector Laboratories (Cat. #H-3502; Newark, California, USA). Images were taken using the Echo Revolve Inverted Microscope (Echo, California, USA).

### Immunofluorescent staining

Immunofluorescent staining was performed to detect the presence of epithelial (EpCAM) and leukocyte markers (CD45). The 3D microtumors were fixed using 4% paraformaldehyde (Cat. #J19943.K2, ThermoFisher Scientific, Massachusetts, USA) for 10 minutes at room temperature followed by three washes with ice-cold PBS. Samples were blocked for 30 minutes using a blocking buffer (3% Human Serum in PBS) at room temperature. Antibody solution containing 1:300 dilutions of AF647 Anti-EpCAM (Cat. #ab237385, RRID: AB_2940927) and AF488 Anti-CD45 (Cat. #ab40763; RRID: AB_726545) antibodies (Abcam, Cambridge, UK) was prepared using the blocking buffer. After 2 hours, samples were washed 3 times with ice-cold PBS. Samples were counterstained with Hoechst 33342 (Cat. #40044; Biotium, California, USA) and imaged using the Echo Revolve Inverted Microscope (Echo, California, USA).

## Data analysis

Four-parameter logistic dose–response curves were fit in GraphPad Prism v10.6.1 using a four-parameter logistic model. DSS was estimated and scaled from 0 (resistant) to 100 (sensitive)^24^. Concordance was determined using Lin’s correlation coefficient (ρc). Agreement between matched primary tumors and metastases, as well as between spatially distinct tumor samples, was assessed using Bland–Altman analysis. Bias was calculated as the mean difference between paired measurements, and limits of agreement were defined as the bias ± 1.96 standard deviations of the differences.

## Data availability

The datasets generated and analyzed during the current study are available from the corresponding author on reasonable request.

## Code availability

No custom code was generated for this study.

## Acknowledgements

We thank the patients and clinical teams who contributed specimens to this study.

## Author contributions

R.N. and I.T. contributed equally. R.N., I.T. and C.A. conceived and designed the study. R.N., I.T., C.M. and R.J.P. performed the experiments and acquired the data. R.N., I.T. and C.M. analyzed and interpreted the data. W.C., B.A. and C.A. provided supervision and resources. R.N. and I.T. drafted the manuscript. All authors reviewed and approved the final manuscript.

## Competing interests

The authors are affiliated with SageMedic Corp., which develops the functional profiling platform described in this study.

## Supplementary information

**Supplementary Fig. S1.**
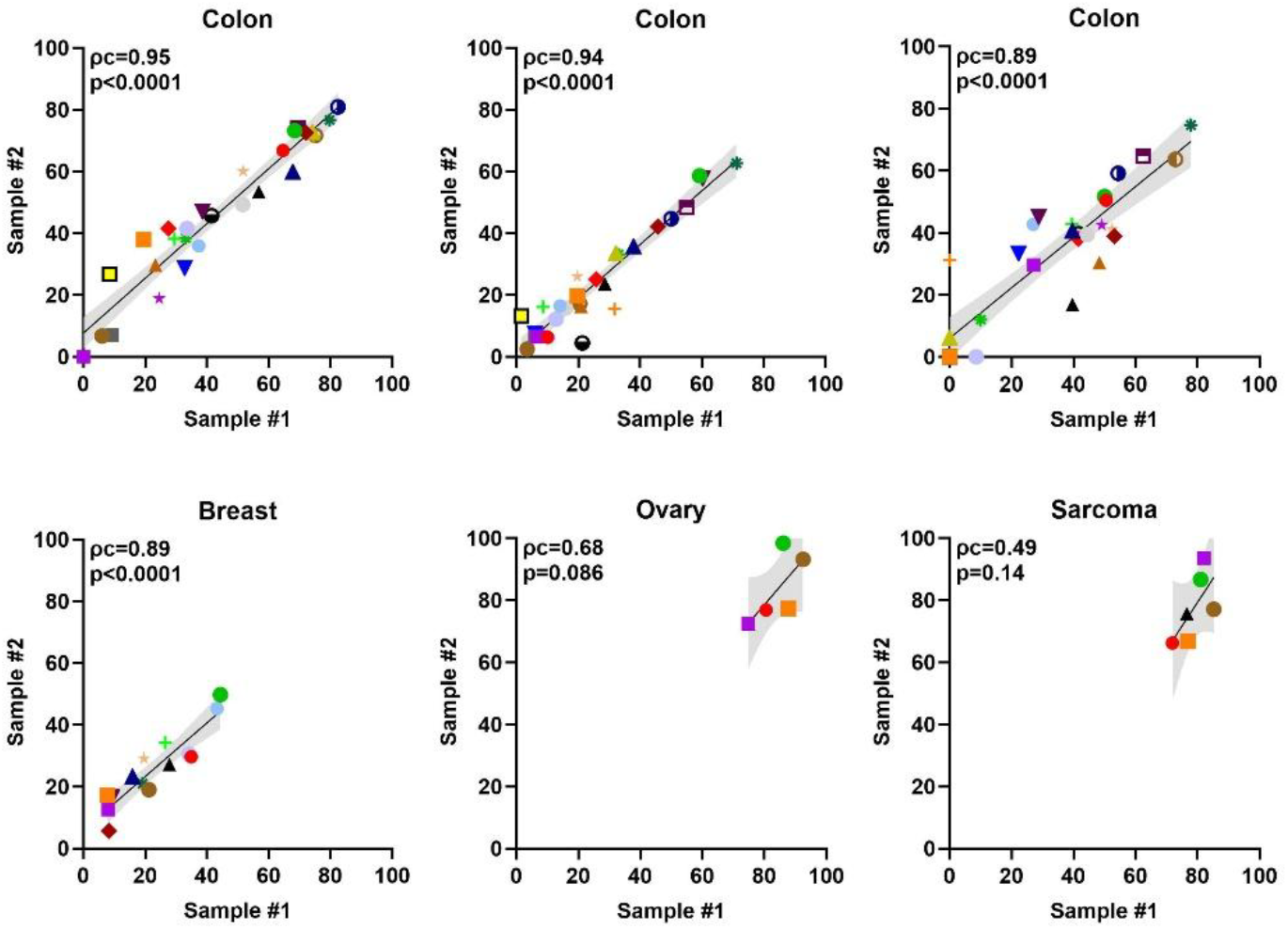
Correlation charts of the effective standardized dose between two distinct locations (sample 1 and 2) of each tumor.

**Supplementary Fig. S2.**
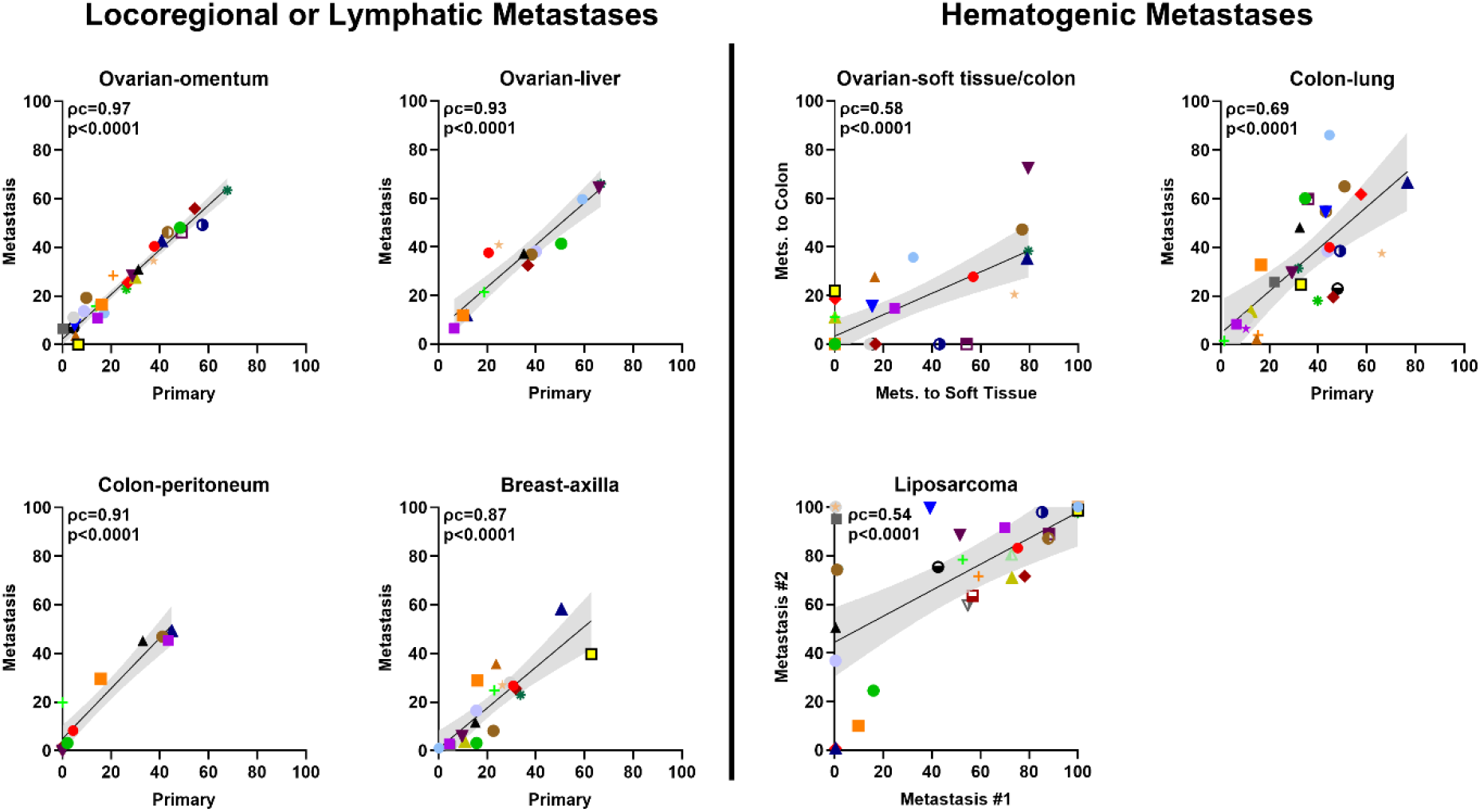
Correlation charts of the effective standardized dose in four pairs of primary tumor and their metastasis. Strong correlation was observed for locoregional metastases of ovarian cancers (ρc = 0.97 and 0.93) and colon cancer (ρc = 0.91), and for lymphatic metastasis of a breast cancer (ρc = 0.87). Weaker correlation was observed for hematogenic metastasis of colon cancer to the lungs (ρc = 0.69), ovarian metastases (ρc = 0.58) and liposarcoma (ρc = 0.54).

**Supplementary Fig. S3.**
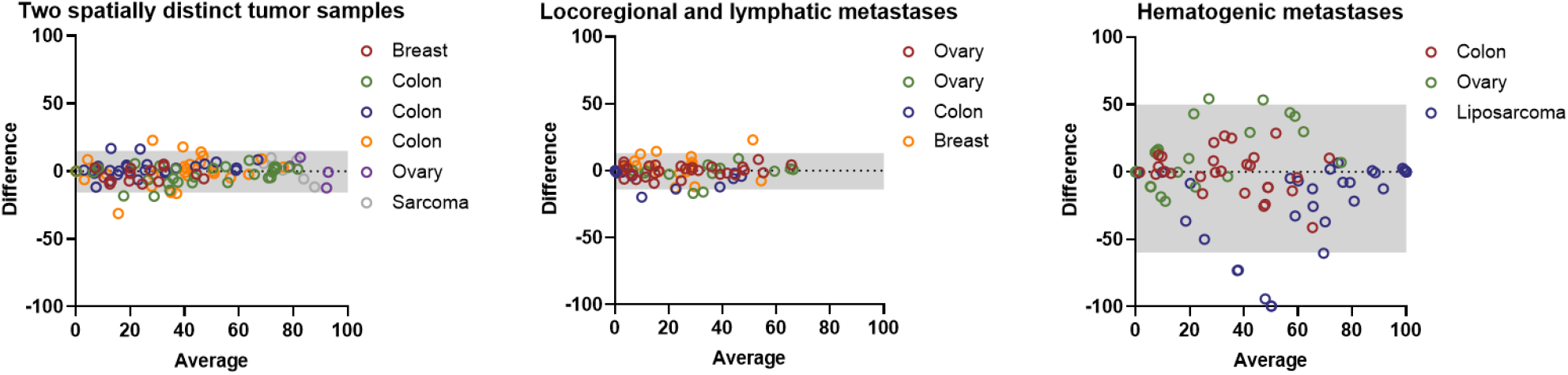
Bland–Altman plots comparing two spatially distinct tumor samples (Fig. 2), locoregional metastases, lymphatic metastases, and hematogenic metastases (Fig. 5). The grey shaded area represents the 95% limits-of-agreement interval.

## Notes

### Author Declarations

The Compassionate Access Institutional Review Board waived the requirement for ethical approval for this work by determining that the research was exempt under 45 CFR 46.104(d)(4)(iii) (IORG0012307; reference CAIRB-26-0901). This determination covered the program Secondary Research Analyses of SAGE Oncotest Results and Associated Clinical Data. The research involved secondary analysis of existing clinical assay results and associated patient data, with patient informed consent and valid HIPAA authorization for research use. No new specimen collection, additional specimen testing, research-specific patient intervention, or treatment assignment was performed.

